# Determining the risk of developing symptomatic covid-19 infection after attending hospital for radiological examinations: controlled cohort study

**DOI:** 10.1101/2021.03.08.21253143

**Authors:** Nikos Evangelou, Sian Vaughan, Aimee Hibbert, Paul S Morgan, Matthijs Backx, Louise Berry, Tim Card, Emma Tallantyre

## Abstract

**OBJECTIVE:** To determine whether brief attendance for outpatient radiological investigations is associated with increased risk of clinically significant coronavirus disease 2019 (covid-19) infection.

**DESIGN:** Observational cohort study with a historical control.

**SETTING:** 2 large UK University Hospitals located in Nottingham and Cardiff.

**PARTICIPANTS:** All 47,340 patients who attended an outpatient radiology appointment at Nottingham University Hospitals and University Hospital of Wales during the first wave of the pandemic in 2020, and 70,655 patients that comprised the control cohort who attended for outpatient radiology the same period in 2019.

**MAIN OUTCOME MEASURES:** The risk of developing clinically significant covid-19 infection within 28-days of attending a radiological examination. Covid-19 infection rates for the 2020 cohort were compared against a control group who attended in 2019.

**RESULTS:** 84 positive SARS-CoV-2 tests were temporally associated with 47,340 radiological examinations across two hospitals in 2020. This low infection rate was higher than the 2019 control cohort; OR 2.507 (1.766 – 3.559) and equates to an approximate 1 positive covid-19 infection per 1000 radiology investigations.

**CONCLUSIONS:** Our data suggests that attending hospitals for outpatient radiological investigations during the pandemic is associated with a very small absolute risk of acquiring clinically significant covid-19 infection. It is unlikely that this risk is directly attributable to radiology attendance, considering the reasons leading individuals to attend hospitals during the pandemic, the true attributable risk will likely be even lower.

**TRIAL REGISTRATION:** ClinicalTrials.gov NCT04544176

## Introduction

Attendance for healthcare appointments for non-covid-19 related diseases has dramatically declined during the coronavirus disease 2019 (covid-19) pandemic. Internationally, hospitals have reduced face-to-face appointments in favour of telephone/video consultations and now follow strict guidelines for enhanced cleaning measurements between face-to-face outpatient consultations and radiology procedures^1^. A reduction in new diagnoses of cancer and stroke, and reduced hospitalisations for acute myocardial infarction has been observed in several studies internationally^2^. Anecdotally, patients’ fear of contracting covid-19 in hospital settings contributes to the decline in attendance for investigations and treatments,^3^ possibly leading to substantial increases in avoidable non-covid-19 morbidity and mortality.^4-8^

Although risk of covid-19 has been reported to be increased during hospital inpatient stays,^9^ the risk of acquiring covid-19 during face-to-face outpatient consultations or radiological investigations has not been reported.

We wanted to investigate whether the risk of acquiring clinically significant covid-19 infection was increased by attending hospitals as an outpatient. We chose to study outpatient attendance for radiological investigations as records allowed reliable verification of in-person attendance. As patients attending hospitals for investigations are more likely to have predisposing comorbidities than the general population, which in itself is a risk factor for covid-19 infection, we compared a 2020 cohort with a control cohort who underwent radiological investigations during an equivalent period in 2019. We hypothesised that a single brief attendance for radiological investigation does pose a small absolute risk of contracting covid-19.

## Methods

This cohort study with a historical control group, examined all patients who attended Nottingham University Hospitals (Nottingham) and the University Hospital of Wales (Cardiff) for outpatient radiological investigations during the first wave of the covid-19 pandemic in the UK. Both study sites are large University NHS hospitals and are the only centres in the cities of Nottingham and Cardiff with an emergency department.

Electronic patient records of hospital attendances for radiological investigations in 2019 and 2020 were extracted by local NHS hospital analysts. All local policies regarding data protection were followed, and all statistical analysis took place at Nottingham University Hospitals, NHS Trust, who sponsored this study. Patients and the public were not involved in the conceptualisation of this research.

SARS-CoV-2 PCR testing was performed at both hospitals and all test results from the onset of the pandemic until 24/5/2020 were available to the research team. During the early phase of the pandemic in the UK and for the duration of this study period, covid-19 testing was offered mainly to symptomatic patients who presented to hospital. All patients hospitalised for suspected covid-19 were also tested.

### Inclusion Criteria

The incubation period for most patients with covid-19 is up to 14 days and modelling suggests that only 1% of patients have a longer incubation period.10 Similarly, 99% of patients who tested positive in the UK did so within 14 days of symptom onset.11 We therefore considered that PCR tests which occurred within the period 28 days after any outpatient radiological investigation would capture most relevant infections.

The first patient identified in Nottingham and potentially the first UK confirmed case with a positive SARS-CoV-2 test was admitted on 21/2/2020.^12^ The first positive SARS-CoV-2 patient in Cardiff was confirmed on 30/3/2020. The exposed cohort comprised all patients who attended an outpatient radiology appointment at NUH between 29/1/2020 and 24/5/2020, and in Cardiff, between 7/3/20 and 11/5/20 who were subsequently tested for covid-19 within 28-days of their attendance. Patients who had an inpatient stay within 28 days of their outpatient radiology examination, but had PCR testing >2 days after admission, were excluded as it was possible that covid-19 infection could relate to the hospital admission rather than outpatient attendance. We included patients admitted to hospital up to 2 days prior to testing. Acquisition of infection in the inpatient setting is expected to have an incubation period exceeding 48 hours, whereas a positive SARS-CoV-2 test within the first 2 days is likely to be community acquired rather than healthcare associated.^13^ Radiological examinations that were part of an inpatient stay were not considered.

An unexposed cohort formed the control group and comprised all individuals who attended outpatient radiological appointments during the same period for each hospital in 2019. These individuals were excluded if they attended radiology during the relevant 28-day risk period in 2020, or if they had an inpatient stay during the same 28-day risk period in 2020.

### Data Validation and Sensitivity Analysis

100 randomly selected participants present in the 2020 datasets extracted by the NHS hospital analysts from Cardiff and Nottingham, were independently validated to confirm that patients attended face-to-face radiology investigations. Conversely, pilot data on outpatient clinic attendances, initially considered for inclusion in the study showed at least 25% of outpatient consultations that were documented to have taken place face-to-face, in fact occurred via telephone and were incorrectly captured on Patient Management Systems. Based on these findings, attendance for radiology outpatient appointments became the focus of this study.

Data verification was also performed for all Nottingham and Cardiff patients who tested positive in 2020 for covid-19. This validated physical in-person radiology attendance and confirmed that our datasets excluded patients who cancelled or did not attend their appointments. This validation also confirmed that attendances were not part of an inpatient stay, and did not occur following a hospital admission in the previous 28 days. Any patients who had an outpatient chest x-ray because of symptoms compatible with covid-19 infection were also excluded.

Accepting that 25% of in-person outpatient consultations were misclassified, we still performed an additional sensitivity analysis on the Nottingham cohort to examine whether outpatient consultations (in addition to radiological appointments) influenced the risk of subsequent covid-19 infection.

As the dates of inclusions for the Nottingham and Cardiff cohorts were different, we also performed analysis on the Nottingham data to mirror the same reporting period from Cardiff: 7/3/20 – 11/5/20. A sensitivity analysis was also performed which included patients who had a covid-19 test up to 5-days following admission.

This article follows the STROBE statement for reporting of cohort studies (https://www.strobe-statement.org/fileadmin/Strobe/uploads/checklists/STROBE_checklist_v4_cohort.pdf).

### Data Analysis

Data analysis was performed using R-4.0.1; the proportion of each cohort experiencing covid-19 infection within the 28-day follow up period was compared via odds ratio. Subjects with positive tests and their respective odds ratios were calculated separately for each hospital.

## Results

47,340 radiological examinations across two hospitals in 2020 were temporally associated with 84 positive SARS-CoV-2 tests; this was compared against 70,655 examinations in 2019 and a respective 50 positive SARS-CoV-2 tests. The rate of receiving a positive test within 28-days of a radiological investigation for the exposed 2020 cohort was higher than the rate of infection during the relevant 28-day window in 2020, for the unexposed 2019 cohort (table 1). 0.07% of those radiologically investigated in 2019 had a positive covid-19 test compared to 0.18% of 2020 participants giving an absolute rate difference of just over 1 covid-19 infection per 1000 investigations. When considering this as the odds of positive test, these were significantly elevated in the 2020 cohort, OR ratio 2.5 (95% CI 1.8-3.6).

**Table 1.** Combined SARS-CoV-2 tests from Cardiff and Nottingham hospitals within 28-days of a single radiological investigation. Time frames: Cardiff (7 March – 11 May), Nottingham (29 January – 24 May).

|  | Nottingham and Cardiff |  |
| --- | --- | --- |
|  | 2019 unexposed cohort | 2020 exposed cohort |
| Total outpatient radiology attendances | 70,655 | 47,340 |
| Outpatient radiology attendances without a covid-19 test | 68,859 | 44,317 |
| Outpatient radiology attendances with a covid-19 test and no associated inpatient stay | 1796 | 3023 |
| Outpatient radiology attendances with a covid-19 test within 28 days of radiology appointment | 431 | 609 |
| Positive covid-19 test | 50 | 84 |
| Negative covid-19 test | 381 | 525 |
| Positive covid-19 test/total outpatient radiology attendances | 0.071% | 0.177% |
| Odds ratio (positive vs number of attendances) | 2.507 (1.766 – 3.559) |  |

We calculated the proportion of positive tests separately for each hospital to assess the differences in risk between the two centres (Table 2).

**Table 2.** SARS-CoV-2 tests within 28-days of a single radiological investigation at the separate centres of Nottingham and Cardiff. Time frames: Cardiff (7 March – 11 May), Nottingham (29 January – 24 May).

|  | Nottingham |  | Cardiff |  |
| --- | --- | --- | --- | --- |
|  | 2019 | 2020 | 2019 | 2020 |
| Total outpatient radiology attendances | 32,495 | 29,903 | 38,160 | 17,437 |
| Outpatient radiology attendances without a covid-19 test | 31,779 | 28,328 | 37,080 | 15,989 |
| Outpatient radiology attendances with a covid-19 test and no associated inpatient stay | 716 | 1,575 | 1,080 | 1,448 |
| Outpatient radiology attendances with a covid-19 test within 28 days of radiology appointment | 124 | 273 | 307 | 336 |
| Positive covid-19 test | 12 | 9 | 38 | 75 |
| Negative covid-19 test | 112 | 264 | 269 | 261 |
| Positive covid-19 test/total outpatient radiology attendances | 0.037% | 0.030% | 0.100% | 0.430% |
| Odds ratio (positive vs number of attendances) | 0.815 (0.34 – 1.93) |  | 4.319 (2.92 – 6.38) |  |

The total number of patients who received a positive SARS-CoV-2 test after radiological investigation in Nottingham in 2020 was 10, while 95 patients tested positive in Cardiff during this period. In the control group, 12 patients in Nottingham received a positive test, compared to 38 patients in Cardiff. Of the patients in 2020 with a positive test, 21 were excluded on the basis that their outpatient investigation was a chest x-ray for symptoms compatible with covid-19 (1 in Nottingham and 20 in Cardiff). The total values included in our analysis for those testing positive in 2020 for Nottingham was 9, compared to 75 patients in Cardiff.

Based on these results, the odds of receiving a positive test following a radiological investigation in 2020 compared to the 2019 cohort was only statistically significantly increased for Cardiff.

As widespread testing was not available in the first month of the Nottingham study period, we examined also the ratio of positive tests to the total number of outpatient radiology examinations undertaken in Nottingham for the same time period as Cardiff. A total of 28,702 patients attended both hospitals for a radiological examination in 2020 compared to 57,311 in 2019. We found that the ratio of positive covid-19 tests per radiological examinations was higher in 2020 but very small (supplementary table 1).

In further sensitivity analysis we compared the differences in risk for Nottingham when the same reporting period of Cardiff was used (7 March - 11 May). A total of 11,265 patients attended outpatient radiology appointments in Nottingham during the 2020 study period and 19,151 during the 2019 period. The positive tests were not more common in the exposed cohort (supplementary table 2).

When for the Nottingham cohort, we excluded patients that attended outpatient clinic appointments between 28 and 2 days prior to a covid-19 test, we only found 4 patients who received a positive test in 2020 and 12 patients in 2019; this ratio was not significantly different from 2019 (supplementary table 3).

To explore whether patients admitted to hospital had a delayed positive test, we also extended the analysis to include also all positive SARS-CoV-2 tests that were detected up to 5 days following admission. The results were not different (supplementary table 4).

## Discussion

Patients who visited hospitals for radiological investigations during the first wave of the pandemic had only a very small risk of acquiring clinically significant covid-19 infection. The absolute increase in risk of covid-19 infection for those attending during the first wave of the pandemic compared to a similar group attending in 2019 was in the region of 1 additional covid-19 infection per 1000 radiology appointments.

Our findings are in keeping with prior findings that the risk of onwards covid-19 transmission is relatively low in outpatient healthcare settings, ^14^ and are reflective of real world experience in an unselected cohort which is more likely to be generalisable to similar populations. Our study does however have some limitations.

In the early phase of the pandemic, covid-19 testing capacity was very small with limited community testing taking place. In our centres, PCR tests were restricted to hospital attendances and admissions. It is therefore possible that patients who attended hospital for outpatient radiology investigations might have subsequently experienced mild or asymptomatic covid-19 infection but were never tested. However, our methodology can be expected to have detected all cases of severe covid-19, requiring hospitalisation, which we feel is most relevant as it seems severe rather than asymptomatic disease drives patient fear of hospital attendance.

It is also likely despite our study design attempting to match to some degree for comorbidity, that our results were influenced by residual cofounding. We suspect individuals attending in 2020, during the pandemic when many patients were avoiding hospitals, were likely to be at higher risk of covid-19 as they had more acute illnesses, with greater comorbidity and were probably exposed to more out of hospital care, compared to the 2019 control cohort. As these factors would increase the risk of severe infection with covid-19, they could lead to over-estimation of the covid-19 risk attributable to a single radiology attendance. The true risk therefore is likely to be below that which we report.

Although we have not found a significantly increased risk of covid-19 infection in Nottingham, a small excess risk was detected in Cardiff. Published government data during our Cardiff reporting period (7/3/20 – 11/5/20) show an increased 7-day average of positive COVID cases in Cardiff (62.7) compared to Nottingham (26.9),^15^ as well as an higher peak number of cases (79) compared to Nottingham (45). As the background rate of covid-19 in Cardiff was therefore higher, the exposure to other Cardiff residents in and out of hospital was likely to be a greater risk than the similar risks in Nottingham.

We have undertaken a number of sensitivity tests to further examine and explain our results. The lack of reduction in Nottingham radiology attendances in the primary analysis is shown to be due to the period studied including a timeframe before widespread concern and behaviour change initiated. When the Nottingham dates were limited to the same reporting frame as Cardiff in supplementary table 2, the figures confirmed this to be true as radiology appointments were more commonly cancelled in Nottingham after March 2020. In addition to try to some degree to exclude the possibility that patients were infected attending outpatient appointments at which radiological investigations were planned in response to, we excluded patients that attended outpatient consultations in addition to outpatient radiological investigations in the Nottingham cohorts. Doing so had little impact upon our results.

Brief attendance in restaurants, bars and coffee shops have all been linked with increased risks,^16^ but it is difficult to disentangle risk attributable to visiting a particular location from overall patient risk-taking behaviour. This is true of attendance for outpatient radiology also. The very small risk of covid-19 infections observed in our study is likely to be influenced by the infection control measures implemented by all UK hospitals, and both staff and the public adhering to social distance guidelines.

Risks of viral exposure will change over time and vary by locality as shown by the differences between our centres. What we observed in Nottingham and Cardiff might be different to other hospitals. However, our results suggest that the hospital attendances for outpatient investigations do not pose a large risk of contracting covid-19 infection. This is reassuring as the benefit of those investigations is likely to far outweigh any potential risk. Since the study period, additional infection control measures have been implemented including mandatory universal mask wearing and encouragement of regular hand sanitisation; these actions may have further reduced the risk of covid-19 acquisition. Hopefully these findings can inform local policy and decision-making by patients, that attendance for a brief hospital appointment is relatively safe.

## Data Availability

Data sharing: A summary of the data may be provided by application to the corresponding author at subject to the necessary ethical and regulatory approvals by the applicant, and subject to data availability.

## Key Findings

1. The absolute risk of covid-19 infection related to outpatient radiology attendance (including the public transport associated with it) was in the region of 1/1000.
2. It is likely that there will be residual confounding influences related to the comorbidities of our study cohorts which will have led to overestimation of the attributable risk.
3. Where community covid-19 infection rates are lower with current safeguards enacted in hospitals, the risk of attendance is negligible.

We thank Matthew Tyler from Cardiff and Vale University Health Board and Craig Hall from Nottingham University Hospitals for their work collecting this data.

## Contributors

NE is the corresponding author. NE, SV, AH, PM, MB, LB, TC and ET conceived and designed the study. SV performed the statistical analysis. NE, AH, TC and ET drafted the manuscript. All authors contributed to acquisition, analysis, or interpretation of data and revised the report and approved the final version before submission.

## Ethical approval

This study was approved by the Health Research Authority (20/HRA/4783) and was not reviewed by a research ethics committee as the research was limited to using previously collected, non-identifiable information.

## Data sharing

A summary of the data may be provided by application to the corresponding author at subject to the necessary ethical and regulatory approvals by the applicant, and subject to data availability.

The lead author (NE) affirms that the manuscript is an honest, accurate, and transparent account of the study being reported; that no important aspects of the study have been omitted; and that any discrepancies from the study as planned (and, if relevant, registered) have been explained.

## Dissemination to participants and related patient and public communities

There are no plans to disseminate the results of the research to study participants. Study results will be shared with the public through the media centre of the authors’ institutions under authors’ supervision.

## Provenance and peer review

Not commissioned; externally peer reviewed.

## Summary Box

### WHAT IS ALREADY KNOWN ON THIS TOPIC

During the coronavirus disease 2019 (covid-19) pandemic, patient attendance to hospitals for non-covid-19 related diseases has declined. Patients’ fear of contracting covid-19 in hospital settings has been shown to contribute to the decline in attendance for investigations and treatments. Although the risk of contracting covid-19 has been reported to increase during hospital inpatient stays, the risk of acquiring covid-19 during face-to-face outpatient consultations or radiological investigations has not been reported.

### WHAT THIS STUDY ADDS

This study found that brief outpatient attendance for radiological investigations carries a small absolute risk of approximately 1 positive covid-19 test for every 1000 outpatient radiological examinations. These findings provide strong real world evidence that the infection control measures implemented in hospitals including enhanced cleaning requirements, mandatory mask wearing and regular hand sanitisation are limiting covid-19 transmission within healthcare settings.

## Supplementary Information

additional tables 1-4.

