## Supplementary Tables for "Determining the risk of developing symptomatic covid-19 infection after attending hospital for radiological examinations: controlled cohort study"

**Supplementary Table 1.**

|  | **Nottingham and Cardiff** | |
| --- | --- | --- |
|  | **2019 unexposed cohort** | **2020 exposed cohort** |
| Total outpatient radiology attendances | 57,311 | 28,702 |
| Outpatient radiology attendances without a covid-19 test | 55,818 | 26,448 |
| Outpatient radiology attendances with a covid-19 test and no associated inpatient stay | 1,493 | 2,254 |
| Outpatient radiology attendances with a covid-19 test within 28 days of radiology appointment | 400 | 553 |
| Positive covid-19 test | 49 | 81 |
| Negative covid-19 test | 351 | 472 |
| Positive covid-19 test/total outpatient radiology attendances | 0.085% | 0.282% |
| Odds ratio (positive vs number of attendances) | 3.301 (2.314 – 4.708) | |

**Supplementary Table 1. Combined SARS-CoV-2 tests from Cardiff and Nottingham hospitals within 28-days of a single radiological investigation.** Time frames: Cardiff (7 March – 11 May), Nottingham (7 March – 11 May).

**Supplementary Table 2.**

|  | **2019 unexposed cohort** | **2020 exposed cohort** |
| --- | --- | --- |
| Total outpatient radiology attendances | 19,151 | 11,265 |
| Outpatient radiology attendances without a covid-19 test | 18,738 | 10,459 |
| Outpatient radiology attendances with a covid-19 test and no associated inpatient stay | 413 | 806 |
| Outpatient radiology attendances with a covid-19 test within 28 days of radiology appointment | 93 | 217 |
| Positive covid-19 test | 11 | 6 |
| Negative covid-19 test | 82 | 211 |
| Positive covid-19 test/total outpatient radiology attendances | 0.057% | 0.053% |
| Odds ratio (positive vs number of attendances) | 0.927 (0.343 – 2.508) | |

**Supplementary Table 2. Nottingham Sensitivity Analysis.** SARS-CoV-2 tests within 28-days of a single radiological investigation at Nottingham hospitals. Same reporting period as Cardiff; 7 March – 11 May for 2019 and 2020.

**Supplementary Table 3.**

|  | **2019 unexposed cohort** | **2020 exposed cohort** |
| --- | --- | --- |
| Total outpatient radiology attendances | 32,327 | 28,493 |
| Outpatient radiology attendances without a covid-19 test | 31,779 | 28,328 |
| Outpatient radiology attendances with a covid-19 test and no associated inpatient or outpatient stay | 548 | 165 |
| Positive covid-19 test | 12 | 4 |
| Negative covid-19 test | 86 | 23 |
| Positive covid-19 test/total outpatient radiology attendances | 0.037% | 0.014% |
| Odds ratio (positive vs number of attendances) | 0.378 (0.122 – 1.17) | |

**Supplementary Table 3. Nottingham Sensitivity Analysis.** SARS-CoV-2 tests within 28-days of a single radiological investigation at Nottingham hospitals with additional outpatient appointments from the previous 28-days excluded from the dataset. Reporting period: 29^th^ Jan – 24^th^ May.

**Supplementary Table 4.**

|  | **2019 unexposed cohort** | **2020 exposed cohort** |
| --- | --- | --- |
| Total outpatient radiology attendances | 32,535 | 29,979 |
| Outpatient radiology attendances without a covid-19 test | 31,779 | 28,328 |
| Outpatient radiology attendances with a covid-19 test and no associated inpatient stay | 756 | 1651 |
| Positive covid-19 test | 12 | 9 |
| Negative covid-19 test | 125 | 286 |
| Positive covid-19 test/total outpatient radiology attendances | 0.037% | 0.030% |
| Odds ratio (positive vs number of attendances) | 0.814 (0.343 – 1.932) | |

**Supplementary Table 4. Nottingham Sensitivity Analysis.** SARS-CoV-2 tests within 28-days of a single radiological investigation at Nottingham hospitals including individuals admitted to hospital and tested up to 5 days following admission. Reporting period: 29^th^ Jan – 24^th^ May.
